# Impact of the COVID-19 epidemic on mortality in rural coastal Kenya

**DOI:** 10.1101/2022.04.06.22273516

**Authors:** M Otiende, A Nyaguara, C Bottomley, D Walumbe, G Mochamah, D Amadi, C Nyundo, EW Kagucia, AO Etyang, IMO Adetifa, E Maitha, E Chondo, E Nzomo, R Aman, M Mwangangi, P Amoth, K Kasera, W Ng’ang’a, E Barasa, B Tsofa, J Mwangangi, P Bejon, A Agweyu, TN Williams, JAG Scott

## Abstract

**Background:** The impact of COVID-19 on all-cause mortality in sub-Saharan Africa remains unknown.

**Methods:** We monitored mortality among 306,000 residents of Kilifi Health and Demographic Surveillance System, Kenya, through four COVID-19 waves from April 2020-September 2021. We calculated expected deaths using negative binomial regression fitted to baseline mortality data (2010-2019) and calculated excess mortality as observed-minus-expected deaths. We excluded deaths in infancy because of under-ascertainment of births during lockdown. In February 2021, after two waves of wild-type COVID-19, adult seroprevalence of anti-SARS-CoV-2 was 25.1%. We predicted COVID-19-attributable deaths as the product of age-specific seroprevalence, population size and global infection fatality ratios (IFR). We examined changes in cause of death by Verbal Autopsy (VA).

**Results:** Between April 2020 and February 2021, we observed 1,000 deaths against 1,012 expected deaths (excess mortality -1.2%, 95% PI -6.6%, 5.8%). Based on SARS-CoV-2 seroprevalence, we predicted 306 COVID-19-attributable deaths (a predicted excess mortality of 30.6%) within this period. Monthly mortality analyses showed a significant excess among adults aged ≥45 years in only two months, July-August 2021, coinciding with the fourth (Delta) wave of COVID-19. By September 2021, overall excess mortality was 3.2% (95% PI -0.6%, 8.1%) and cumulative excess mortality risk was 18.7/100,000. By VA, there was a transient reduction in deaths attributable to acute respiratory infections in 2020.

**Conclusions:** Normal mortality rates during extensive transmission of wild-type SARS-CoV-2 through February 2021 suggests that the IFR for this variant is lower in Kenya than elsewhere. We found excess mortality associated with the Delta variant but the cumulative excess mortality risk remains low in coastal Kenya compared to global estimates.

## INTRODUCTION

By December 2021, the global mortality attributable to COVID-19 was more than 5.4 million^1^. In Europe and South America, there were 204 and 274 COVID-19 deaths per 100,000 population, respectively, but in Africa the figure was only 16.6 per 100,000^1^. Four factors may be contributing to this substantially lower recorded mortality: slower spread of infection; younger population structure; under-ascertainment of COVID-19 deaths; or a lower infection fatality ratio (IFR)^2^. Seroprevalence estimates for SARS-CoV-2 antibodies are now greater than 50% in several African countries^3-10^, suggesting widespread exposure to infection. The younger population structure could reduce the population mortality attributable to COVID-19 but it is insufficient to account for the very low numbers of COVID-19 deaths detected in Africa^11^. Analyses of ascertainment bias and IFR require observations of mortality independent of routine COVID-19 case detection systems.

Excess mortality is an alternative measure of the impact of COVID-19 that is independent of testing availability and has been used in several countries to evaluate the death toll of COVID-19^12-15^. Modelled estimates of excess mortality due to COVID-19 suggest an excess of 120 deaths per 100,000 population globally and an excess of 102 per 100,000 in sub-Saharan Africa^16^. Estimating excess mortality in Africa is challenging because only 8 out of 54 countries have comprehensive Civil Registration and Vital Statistics Systems (CRVS)^17,18^. By the end of 2021, South Africa, Egypt, Mauritius, Tunisia and Seychelles, all of which have complete CVRS, reported excess deaths of 406, 265, 86, 44 and -117 deaths per 100,000 population, respectively^1^. The estimates vary widely and may not generalize well to continental sub-Saharan Africa. Among countries without adequate CRVS, mortality data are usually acquired through decennial censuses, 3-5 yearly cluster sample surveys (Demographic and Health Surveys) or open cohort studies (Health and Demographic Surveillance Systems). Here, we use mortality data from the Kilifi Health and Demographic Surveillance System (KHDSS), a surveillance population of over 300,000 residents followed for 20 years, to examine the mortality impact of COVID-19 in an area typical of rural Kenya.

In a random population survey in the KHDSS between December 2020 and April 2021, 14.5% of children (0-15 years) and 25.1% of adults (≥16 years) were anti-SARS-Cov-2 IgG positive, suggesting widespread transmission by early 2021^19^. We examine mortality trends up to 23^rd^ September 2021 and investigate the impact of the pandemic on morbidity using a hospital surveillance system linked to the KHDSS.

## METHODS

### Data source and study setting

The first case of COVID-19 in Kenya was identified on 12^th^ March 2020 and 295,028 cases and 5,378 deaths were reported up to the end of December 2021^1^. There have been five waves of COVID-19 in Kenya up to March 2022. The fourth (Delta) wave peaked in August 2021 (Figure S1) within our study period.

The KHDSS was established in 2000^20^ with an initial census of 180,000 residents. Vital status and migration events have been recorded in subsequent re-enumeration rounds conducted every 4 months. By 2020, the population size was 306,000 and approximately 1300 deaths were recorded in each of the three years 2017-19^21^. In KHDSS, the first case of COVID-19 was detected on 22^nd^ March 2020.

Cause of death within the KHDSS has been evaluated by Verbal Autopsy (VA) since 2008^22^. A trained field interviewer asks close relatives about signs and symptoms of the deceased household member from 30 days after the date of death. The interviews use the 2014 WHO VA questionnaire and are coded by the InterVA-4 algorithm^23^. From April 2020 we asked 6 additional questions (Table S1) based on WHO recommendations to identify possible COVID-19 deaths^23^. Because of a nationwide lockdown, KHDSS field operations were suspended between 23^rd^ March 2020 and 25^th^ October 2020.

Kilifi County Hospital (KCH) is a first-level hospital located centrally within the KHDSS. It has 66 adult beds and 14 beds in an amenity ward which served as a temporary isolation unit. There are few inpatient beds in other health facilities serving this population. Admissions to KCH are linked in real-time to the KHDSS population register to provide a hospital-based passive disease surveillance system.

We restricted morbidity analysis to adults aged 15 years or older and analyzed the patterns of clinical pneumonia admissions and all admissions. Clinical pneumonia, an existing admission diagnosis on our surveillance system, was defined as the presence of any two of the following symptoms within the 14 days before admission: cough, fever, chest pain, crackles, haemoptysis and dyspnoea.

### Statistical analysis

Rates of mortality and hospital admissions were calculated as the number of deaths or admissions divided by person years of observation (PYO). PYO were calculated as time from the latest of birth or in-migration or study start date to the earliest of study end date or outmigration or death. Individuals’ periods of residence outside the KHDSS area were excluded. Models fitted to rates of mortality, all admissions and pneumonia admissions in 2010-2019 were used to predict these outcomes in 2020-2021. We chose January 2010 to December 2019 as the baseline period because mortality rates were stable during this decade^21^. We censored all analyses on 23^rd^ September 2021, the date our 50^th^ KHDSS re-enumeration round was completed, and we locked our demographic database on 14^th^ January 2022, the date of completion of the 51^st^ round (Figure S1).

We modelled monthly death counts using negative binomial regression. The model included a log-linear trend, sine and cosine terms to account for seasonality, and an offset to account for changes in person years of observation (Equation S1). It was fitted using the glm.nb function in R.

We modelled the rates of monthly admissions as a local-level time series model^24^ to account for the stochastic trend of admission rates (Equation S2). The model was fitted, without covariates, to pre-pandemic hospital admissions. The months January-December 2017 were excluded as missing because of a prolonged healthcare workers’ strike. Fitting was done using the Arima function in R; the local level model is equivalent to an ARIMA (0,1,1) model.

We modelled the log rate of pneumonia admissions using a linear regression model with ARMA errors to account for autocorrelation. Because there were zero admissions in some months, we used quarterly rates of admission. The model included a linear trend term, but no terms for seasonality (Equation S3). To fit the model, we used the Arima function in R.

Excess mortality was calculated as the difference between the observed and predicted number of deaths in the COVID-19 period expressed as a percentage of predicted number of deaths or as a risk based on the initial population size (per capita excess mortality). We used 1st April 2020 as the start of the analysis period because the interval from infection to death is approximately 2-3 weeks and it is unlikely there would have been any COVID-19 deaths before April 2020. We analysed two periods of cumulative excess mortality; in the first, we censored deaths and PYO at 17^th^ February 2021, the reference date for the KHDSS serosurvey of SARS-CoV-2 antibodies; in the second, we censored at 23^rd^ September 2021. In each analysis we included any events captured on or before 14^th^ January 2022.

In the last 10 re-enumeration rounds, 99% of deaths were ascertained by the end of the first complete re-enumeration round after the death (Table S2). The remaining 1% were not detected until a second interview was undertaken because the initial respondent, who may have been a member of an adjacent household, may not have been aware of the death. To adjust for this under-ascertainment bias we divided the number of observed deaths occurring between 4^th^ May and 23^rd^ September 2021 (Figure S1) by 0.99.

The KHDSS population is re-enumerated every 4 months because infant deaths may be missed with longer intervals; a child may be born and die without any enumeration contact. The suspension of the KHDSS field operations on 23^rd^ March 2020 extended the re-enumeration interval to 11 months and it is likely that the ascertainment of infant deaths was reduced in this period. Therefore, we excluded infants from the estimation of excess mortality. In-migrants may also enter and die before they can be enumerated. To exclude mortality changes attributable to variable detection of in-migrants, we compared the annual mortality risk of the cohort of KHDSS residents on 23^rd^ March in 2020 to similar snapshot cohorts on 23^rd^ March in each year of the baseline period.

Expected COVID-19 deaths were calculated as the product of the global IFR for SARS-CoV-2 infection^25^, the cumulative incidence of SARS-CoV-2 and the number of KHDSS residents in each age stratum. We calculated confidence intervals for the expected number of COVID-19 deaths using the delta method^26^.

The cumulative incidence of SARS-CoV-2 was derived from a serosurvey of a random sample of 850 KHDSS residents between 1^st^ December 2020 and 28^th^ April 2021 (median sample date 17^th^ Feb 2021). The serosurvey sampled 100 individuals in each 5-year age stratum from 0-14 years, 50 individuals in each 5-year age stratum between 15-64 years and 50 individuals aged 65 or greater^19^. We compared deaths predicted by this calculation with the total number of excess deaths observed in all KHDSS residents aged 1 year or more between 1^st^ April 2020 and 17^th^ February 2021.

We calculated cause-specific mortality fractions in 10 categories; the 9 leading causes of death in children and adults, namely, Acute Respiratory Infections (ARI), unspecified cardiac disease, stroke, pulmonary tuberculosis, malaria, HIV-related deaths, digestive neoplasm, acute abdominal conditions, and Road Traffic Accidents (RTA), and all other causes. To identify COVID-19-specific deaths we used the 6 additional VA COVID-19 questions; for any death with at least one positive response, we applied two discriminating processes. Firstly, we invited two independent reviewers to conduct Physician Certified Verbal Autopsy (PCVA) using clinical information collected during VA interview and classified each death as a probable, possible or unlikely COVID-19 death. Discordant cases were resolved jointly by the reviewers. Secondly, we used the COVID-19 Rapid Mortality Surveillance (CRMS) software^27^, which is a simplified version of the probabilistic modelling methods used in the InterVA-4 models, to derive the probability that the death was COVID-19 related. We used a probability cut-off value of 0.89 based on a validation study conducted in Brazil^28^.

## RESULTS

We analyzed 15,379 deaths and 12,295 hospital admissions between 1^st^ Jan 2010 - 23^rd^ September 2021 with a total of 3,157,610 PYO.

Overall hospital admission rates declined markedly across all age groups between 2010-2012 from 13 admissions per 1000 PYO to 8 admissions per 1000 PYO. From 2013, admission rates increased before leveling off in 2015-2019 (Figure 1A-D). During the pandemic period, the observed rate of all admissions was generally lower than the predicted rates in all age subgroups.

**Figure 1.**
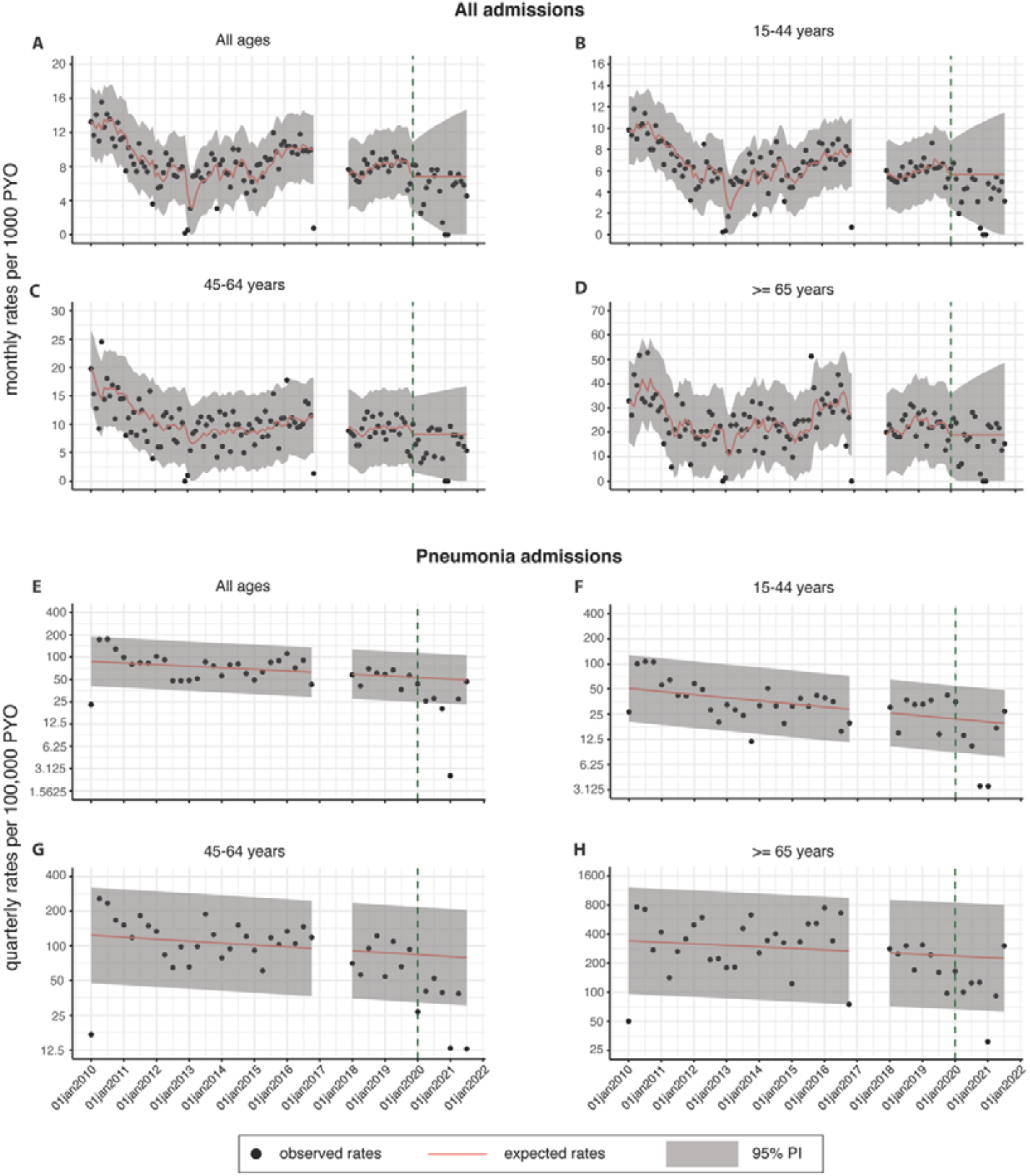
Observed vs expected number of admissions to Kilifi County Hospital with all diagnoses (4A-D) and (pneumonia 4E-H), by age stratum, from 01 April 2020 - 23 September 2021. Data from the year 2017 were excluded as unreliable because there was a year-long strike among health-care workers. Only datapoints prior to January 2020 (dashed vertical line) contributed to the models; expected rates and 95% prediction intervals (PI) were extrapolated from the model to the period January 2020-September 2021.

Pneumonia admission rates declined in the pre-pandemic period across all age groups (Figure 1E-H). For all adults, the incidence declined from approximately 100 cases per 100,000 PYO in 2010 to approximately 50 cases per 100,000 PYO at the end of 2019 (Figure 1E). As with all admissions, the observed rate of pneumonia admissions during the pandemic was lower than the predicted rates in all age subgroups.

Monthly observed mortality rates did not vary consistently from the predicted rates throughout the pandemic, except in July and August 2021 where there was a significant excess of deaths in adults aged ≥45 years (Figure 2). There was marked negative excess mortality in infants throughout the period June 2020-May 2021 suggesting under-ascertainment of infant deaths (Figure 3). Beyond infancy, there was negative excess mortality in most months between April 2020 and May 2021. The marked excess mortality in July and August 2021 was attributable to deaths in persons aged 45-64 years and ≥65 years (Figure 3).

**Figure 2.**
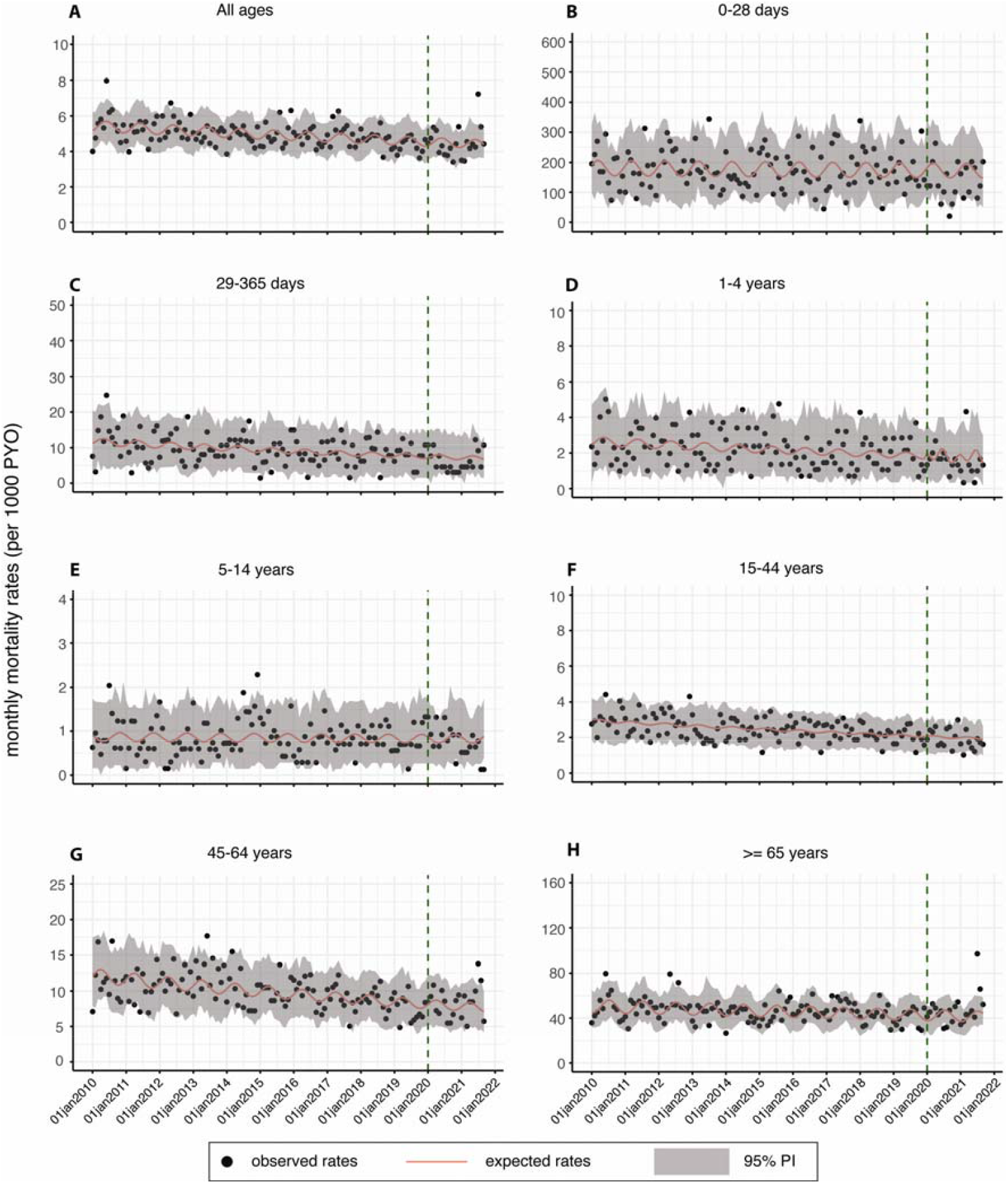
Observed and expected death rates from 01 April 2020 - 23 September 2021. Expected deaths rates in 2020 and 2021 were estimated from a negative binomial regression fitted using data up to 2019

**Figure 3.**
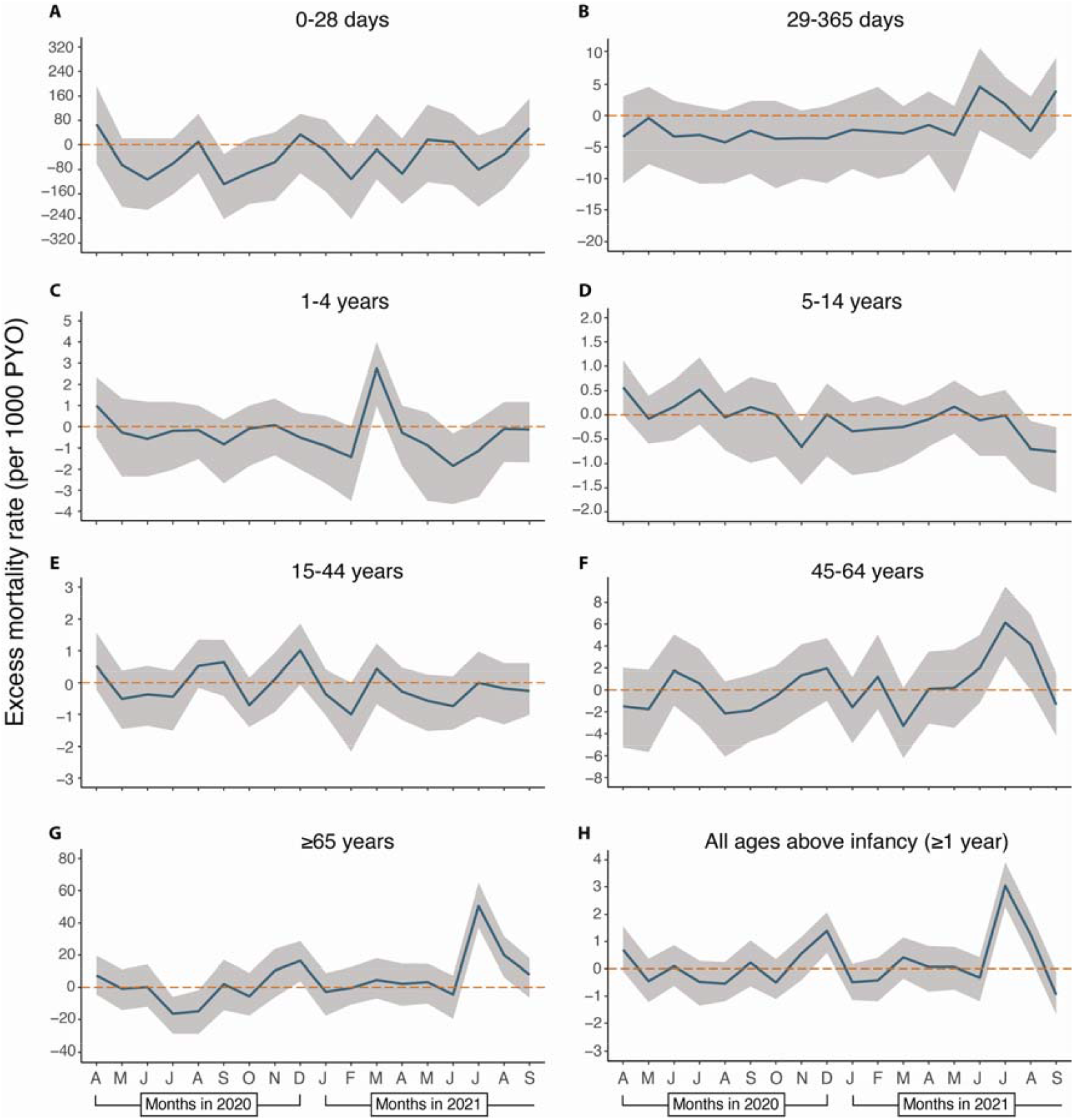
Monthly excess mortality rates from 1^st^ April 2020 – 23^rd^ September 2021. Calculated as (observed deaths - expected deaths)/PYO. Excess mortality rate for all ages above infancy (≥ 1 year) is the weighted average of the age-specific rates. The weights are the proportion of each age-group in the KHDSS population.

Between 1^st^ April 2020 and 17^th^ February 2021, we predicted 1,012 deaths beyond infancy but observed 1,000 (excess mortality -1.2%, 95% PI -6.6%, 5.8%, Table 1). Excess mortality was significantly negative among neonates (−25.8% 95% PI -37.5, - 1.5%) and older infants (−38.0%, 95% PI -48.8, -16.5%, Table S3) up to 17^th^ February 2021. Between 1^st^ April 2020 and 23^rd^ September 2021, we predicted 1,718 deaths beyond infancy but observed 1,768 (Table 1); this gives an excess mortality of 2.9% (95% PI -1.0%, 7.8%) and excess mortality risk of 16.8/100,000. After adjusting for under-ascertainment of deaths occurring between 4^th^ May-23^rd^ September, excess mortality was 3.2% (95% PI -0.6%, 8.1%) and excess mortality risk was 18.7/100,000 (Table S5). The adjusted excess mortality deviated significantly from zero only among adults aged ≥65 years (11.0% 95% PI 5.0%, 19.8%).

**Table 1.**
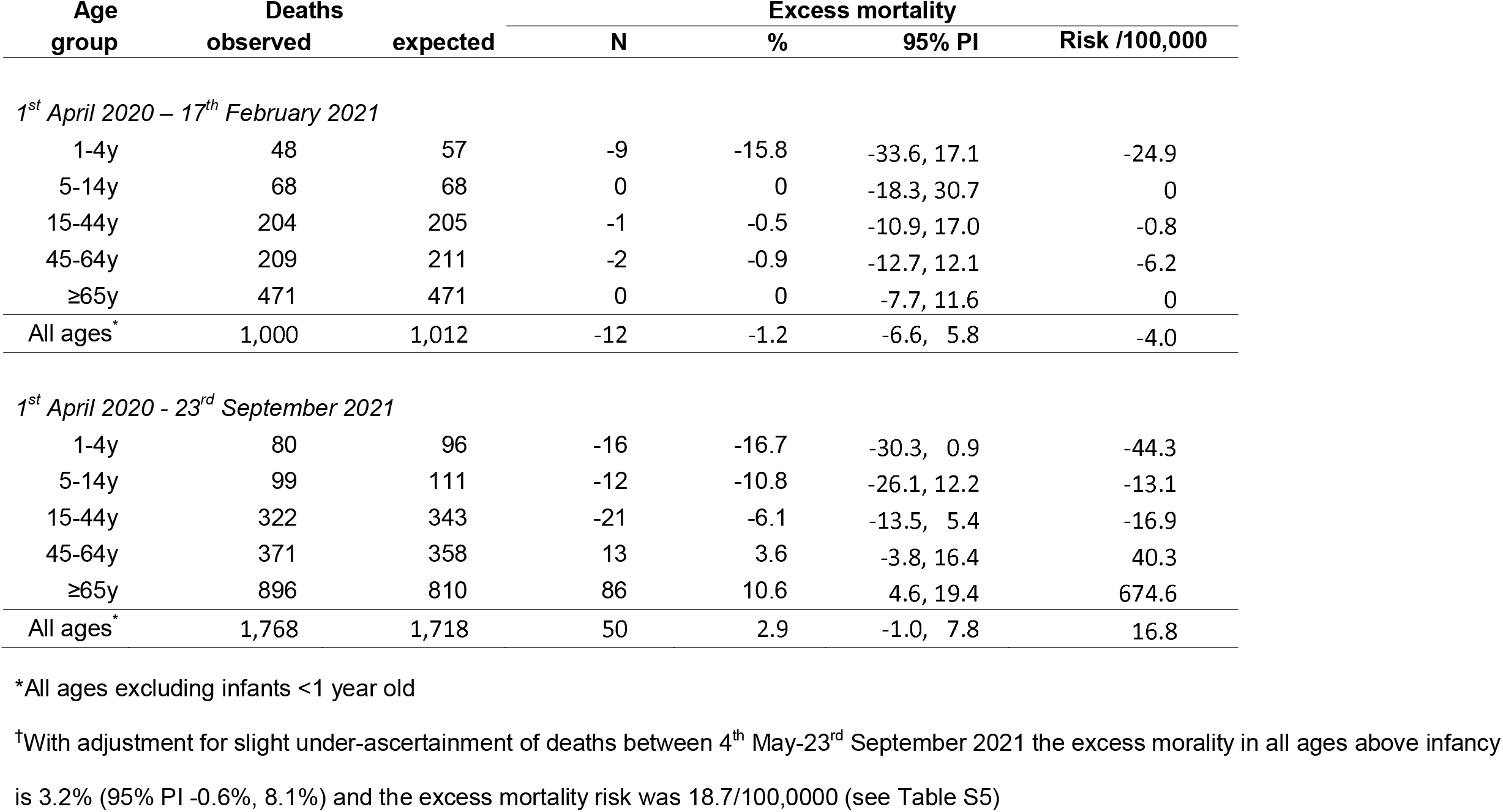
Excess deaths 1st April 2020-17th February 2021 and 1st April 2020-23rd September 2021 in KHDSS residents aged ≥1y

| Age group | Deaths |  | Excess mortality |  |  |  |
| --- | --- | --- | --- | --- | --- | --- |
|  | observed | expected | N | % | 95% PI | Risk /100,000 |
| *1<sup>st</sup> April 2020 – 17<sup>th</sup> February 2021* |  |  |  |  |  |  |
| 1-4y | 48 | 57 | -9 | -15.8 | -33.6, 17.1 | -24.9 |
| 5-14y | 68 | 68 | 0 | 0 | -18.3, 30.7 | 0 |
| 15-44y | 204 | 205 | -1 | -0.5 | -10.9, 17.0 | -0.8 |
| 45-64y | 209 | 211 | -2 | -0.9 | -12.7, 12.1 | -6.2 |
| ≥65y | 471 | 471 | 0 | 0 | -7.7, 11.6 | 0 |
| All ages* | 1,000 | 1,012 | -12 | -1.2 | -6.6, 5.8 | -4.0 |
| *1<sup>st</sup> April 2020 - 23<sup>rd</sup> September 2021* |  |  |  |  |  |  |
| 1-4y | 80 | 96 | -16 | -16.7 | -30.3, 0.9 | -44.3 |
| 5-14y | 99 | 111 | -12 | -10.8 | -26.1, 12.2 | -13.1 |
| 15-44y | 322 | 343 | -21 | -6.1 | -13.5, 5.4 | -16.9 |
| 45-64y | 371 | 358 | 13 | 3.6 | -3.8, 16.4 | 40.3 |
| ≥65y | 896 | 810 | 86 | 10.6 | 4.6, 19.4 | 674.6 |
| All ages* | 1,768 | 1,718 | 50 | 2.9 | -1.0, 7.8 | 16.8 |

\*All ages excluding infants <1 year old
†With adjustment for slight under-ascertainment of deaths between 4<sup>th</sup> May-23<sup>rd</sup> September 2021 the excess mortality in all ages above infancy is 3.2% (95% PI -0.6%, 8.1%) and the excess mortality risk was 18.7/100,000 (see Table S5)

Based on the SARS-CoV-2 seroprevalence estimates and the global IFR, the cumulative number of expected COVID-19 related deaths up to 17^th^ February 2021 was 306 (95% CI, 249-376, Table 2) which represents a total excess mortality of 30.6%. This model did not predict any deaths in infancy and, within this same period, we observed 12 fewer deaths than predicted, giving a negative excess mortality of - 1.2%.

**Table 2.** Expected COVID-19 related deaths up to 17^th^ February 2021 calculated using seroprevalence estimates as an indicator of cumulative incidence and Levin’s estimates for the IFR of SARS-CoV-2 infection

| Age group (years) | KHDSS |  |  | IFR (Levin) |  |  | Expected KHDSS deaths |  |  |
| --- | --- | --- | --- | --- | --- | --- | --- | --- | --- |
|  | Pop'n size | Sero-prevalence | (95% CI) | Estimate (per 100,000 population) | lower 95% CI | upper 95% CI | estimate | lower 95% CI | upper 95% CI |
| 0-9 | 88,841 | 8.4 | (3.9 - 12.9) | 1 | 0.7 | 1.3 | 0 | 0 | 0 |
| 10-19 | 79,931 | 22.0 | (14.8 - 29.2) | 3 | 2 | 4 | 1 | 0 | 1 |
| 20-29 | 47,468 | 23.2 | (14.9 - 31.6) | 11 | 9 | 13 | 1 | 1 | 2 |
| 30-39 | 32,452 | 32.0 | (22.4 - 41.6) | 37 | 31 | 43 | 4 | 3 | 5 |
| 40-49 | 24,285 | 28.5 | (19.4 - 37.6) | 123 | 108 | 141 | 9 | 7 | 10 |
| 50-59 | 15,298 | 24.1 | (14.8 - 33.5) | 413 | 362 | 471 | 15 | 13 | 17 |
| 60-69 | 11,757 | 14.3 | (6.7 - 22.0) | 1,381 | 1,190 | 1,610 | 23 | 20 | 27 |
| 70-79 | 5,873 | 41.1 | (28.1 - 54.1) | 4,620 | 3,830 | 5,570 | 112 | 92 | 135 |
| 80+ | 2,684 | 34.1 | (0.0 - 70.8) | 15,460 | 12,200 | 19,500 | 141 | 111 | 180 |
| Total |  |  |  |  |  |  | 306 | 249 | 376 |

There was no evidence that field interviewers were unable to reach appropriate respondents during the pandemic leading to under-reporting of deaths (Table S6). In annual period survival analyses for the KHDSS cohorts that were resident on 23^rd^ March each year between 2010 and 2020, there was no evidence of decreased survival in 2020 (Table S7, Figure S5).

Between 2015-2019, 71.7 of the deaths detected in the KHDSS were investigated by verbal autopsy (Figure S2). Between 1^st^ January 2020 and 3^rd^ May 2021 (end of re-enumeration round 49) we investigated 68% (1,152/1,695) of recorded deaths. The proportion attributable to ARI fell in 2020 in all age groups but returned to pre-COVID-19 levels in the first 4 months of 2021 (Figure 4). The proportion attributable to RTA rose throughout the pandemic. Of 72 verbal autopsies where at least one COVID-19 specific answer was positive, 18 were defined as COVID-19 deaths using the algorithm, representing 2% of 911 deaths investigated between 1^st^ April 2020 and 3^rd^ May 2021. By physician review, 9 were probably related to COVID-19, 5 were possibly related and 58 were unrelated (Figure S3).

**Figure 4.**
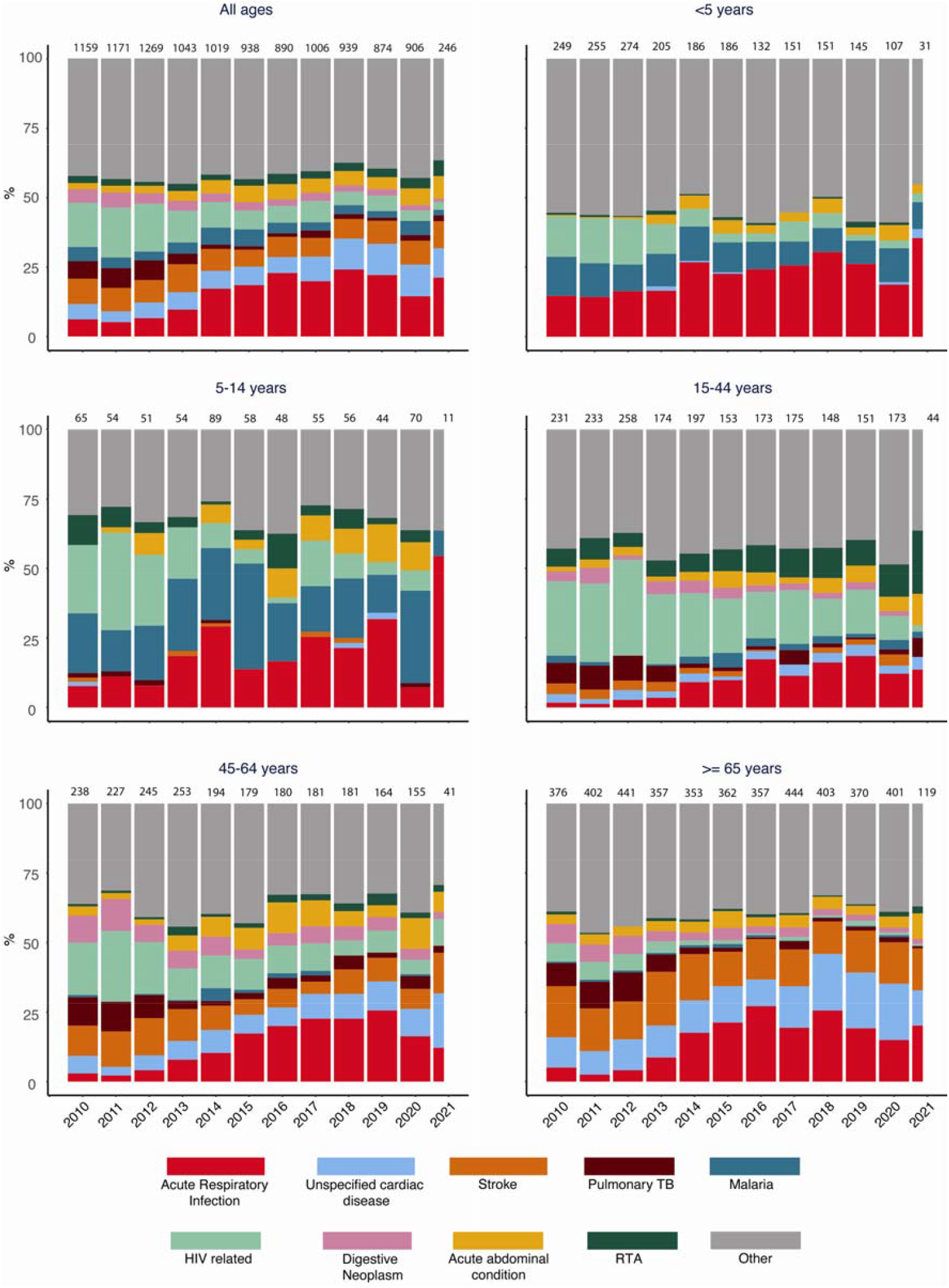
Cause-specific mortality fractions by Verbal Autopsy

## DISCUSSION

We have analyzed hospital admissions, community deaths and the seroprevalence of SARS-CoV-2^19^ within a single large HDSS population in rural Kenya during the first 18 months of the COVID-19 pandemic. Despite two waves of COVID-19 wild-type infections over 11 months and serological evidence indicating SARS-CoV-2 infection in 25.1% of adults and 14.5% of children in the same population, we were unable to identify any signal of increased hospital utilization, either for pneumonia or all other diagnoses combined, in the principal public hospital serving this population. In addition, we did not find any signal of excess mortality in the community during this period. Applying age-specific IFRs from a global meta-analysis of COVID-19 to the cumulative incidence of SARS-CoV-2 and the population structure of KHDSS we would have expected an excess of 306 (30.6%) deaths but, in fact, observed a deficit of 12 (−1.2%) deaths.

It is only in the fourth, Delta wave of COVID-19 that we saw a mortality increase in the KHDSS leading to an excess mortality of 11% in those ≥65 years old. The overall population estimate for excess mortality of 3.2% (95% PI -0.6, 8.1%) obscures considerable heterogeneity, with negative estimates in all age groups below 45 years, and it does not exclude the possibility of zero excess mortality. The population estimate of excess mortality risk attributable to COVID-19 of 18.7/100,000 contrasts sharply with the excess mortality risk estimate of 181.2/100,000 for Kenya as a whole, derived from global mortality models^16^. These estimates differ in scope; (i) the KHDSS population is largely rural and COVID-19 mortality is higher in urban centres^29^; (ii) the observed KHDSS data extend to September 2021 whilst the modelled risks extend to December 2021 and include the tail of the Delta wave and the start of the Omicron wave. Nonetheless, these factors are unlikely to explain the ten-fold difference in estimates which highlights the uncertainty in modelling mortality in Africa without local mortality data.

Among adult residents of the KHDSS area, we observed a sharp fall in all admissions to hospital in March 2020 reflecting the rapid imposition of travel restrictions at the start of the pandemic^30^. However, admissions returned to normal levels within 3 months, reflecting national trends in mobility, captured by Google Community Mobility Reports, which had normalised by October 2020^31^. The incidence of pneumonia admissions was lower than predicted throughout the pandemic period in all age groups. Collectively, these hospital surveillance data did not reveal an epidemic signal of undetected COVID-19 admissions.

The Kilifi County Health Department established a rapid response team to isolate suspected and PCR-confirmed SARS-CoV-2 individuals at the hospital amenity ward and in two external isolation centers. The isolation centers processed a total of 66 suspected COVID-19 cases during their three-month lifespan while the hospital-based isolation ward admitted 139 suspected COVID-19 patients up to May 2021. The number of patients diverted from the KCH general wards to these isolation facilities was small, not all of them were residents of the KHDSS, and even after accounting for these additional hospital admissions we did not observe a signal of increased hospital utilization (Figure S4).

In several countries, the pandemic has led to an increase in deaths at home because patients have been unable or unwilling to travel to hospital for fear of acquiring SARS-CoV-2^32^. It is possible that COVID-19 patients, resident in the KHDSS area, stayed at home for similar reasons. In Kilifi, the excess mortality analysis showed no evidence of an increase in deaths among adults during the first 11 months of the pandemic. Because of the disruption to fieldwork in 2020, infants who were born and died between household visits may not have been reported to the field interviewers. We found a 30% reduction in infant mortality between 1^st^ April 2020 and 17^th^ February 2021 (Table S3) which is best explained by this under-ascertainment and we therefore excluded infants from our analyses of overall mortality. This problem is unlikely in resident children aged 1 year or more, whose births had already been captured by the demographic surveillance before the disruption. Similarly, unhealthy in-migrants, who migrated and died in the period when fieldwork was suspended, may not have been reported. Our mortality analysis excluding in-migrant deaths did not reveal a reduction in survival in 2020 compared to previous years (Table S7, Figure S5).

In the verbal autopsy data, undetected COVID-19 deaths are likely to be identified as Acute Respiratory Infections (ARI). However, the proportion of all deaths attributable to ARI declined in the first year of the pandemic in all adult age groups and then returned to baseline levels in 2021 for most age groups. Among children aged <5 years, ARI declined as a cause of death from 26% in 2019 to 19% in 2020 and rose again in early 2021 consistent with a transient decline in social contacts and respiratory infections during COVID restrictions^33^. Among adults, we also used the VA data to explore the possibility that public health measures reduced mortality from non-COVID-19 causes and this may have masked an excess of deaths caused by COVID-19. For example, reduced travel by road may have led to a reduction in road traffic accident (RTA) deaths. In fact, the proportion of deaths attributable to RTA increased year on year between 2019 and 2021 among those aged 15-44 years old, in whom this is a significant cause of death. The COVID-19 specific VA questions proposed by WHO attributed only 2% of all deaths to COVID-19 from 1^st^ April 2020 to 3^rd^ May 2021. The verbal autopsy data, which do not include the Delta wave, show no indication of a change in cause of death towards categories that would capture COVID-19.

The estimate of cumulative incidence of SARS-CoV-2 in the first 11 months of the pandemic is highly dependent on the accuracy of ELISA for SARS-CoV-2 IgG. This assay, validated in Kenyan control populations, has a specificity of 99.0% (95% CI 98.1-99.5%) and sensitivity of 92.7% (95% CI 87.9-96.1%)^34^. The assay also has 97% agreement with the WANTAI total IgG ELISA adopted by WHO across Africa^35^. The prediction of 306 COVID-19 related deaths in KHDSS derived from the seroprevalence results relies heavily on estimates in older persons yet seroprevalence was estimated in only 101 adults in the three strata above 60 years, of whom 24 were seropositive. Collapsing these age strata and applying a seroprevalence of 24% to all adults aged ≥60 years would lead to a prediction of 234 deaths. Although lower, this still represents a substantial mortality excess which contrasts sharply with the observation of 12 fewer deaths in the same period, suggesting that the IFR of the wild-type virus in Kilifi was substantially lower than was estimated globally^7^.

Excess mortality in South Africa and North Africa^13,36^ has been high during the pandemic period but mortality data from countries in other parts of Africa are sparse. Given seroprevalence estimates of 50%^37^, 60% and higher^38,39^ in many settings in tropical Africa, the lack of a corresponding excess in detectable mortality raises questions about the severity of COVID-19 in this context. Although early waves of the pandemic did not result in an observable risk in mortality in Kilifi, we did observe a discrete peak of mortality coincident with the national Delta wave confined to adults aged ≥45 years. Normally, mortality reporting is subject to lags. To report Delta-wave mortality promptly we have adjusted observed estimates using ascertainment levels derived from many rounds of prior re-enumeration. This yields an excess mortality of 3.2% and an excess mortality risk of 18.7/100,000 across the first four waves of COVID-19 which compares with a global excess mortality risk of 120.3/100,000^16^. Taken together with hospital surveillance showing lower utilization during the pandemic and serological data confirming extensive spread of SARS-CoV-2 infection, these data confirm that the public health impact of the COVID-19 pandemic in coastal Kenya in 2020-21 has been substantially lower than elsewhere in the world.

## Data Availability

Underlying individual data include geo-located residence and individual hospital records and hence would be high risk for identifiability. Intermediary data will be published on the Havard dataverse server under a CCBY 4.0 license.

## Declarations

### Ethics approval and consent to participate

Individual verbal consent to participate in a continuous health and demographic surveillance system was sought at the household level using a specific informed consent form. Written informed consent was obtained by interviewers from all VA respondents. This study was approved by the Ethical Review Committee of the Kenya Medical Research Institute (approval number: KEMRI/SERU/CGMR-C/007/3057).

### Authors’ contributions

Conceptualization: JAGS, TNW, PB. Data collection and preparation: AN, DW, GM, DA, CN. Formal analysis: MO, CB, JAGS Interpretation: MO, JAGS, CB, EWK, AOE, AIMO, EB, BT, JM, PB, AA, TNW. Writing, original draft preparation: MO, JAGS, CB. Writing, reviewing, and editing: all authors. Resources and funding acquisition: PB, JM, BT, JAGS, TNW. All authors critically reviewed the article and approved the final version for submission.

## Acknowledgements

We gratefully thank the residents of Kilifi who have participated in the surveillance activities of the KHDSS. We acknowledge the tremendous work of the verbal autopsy and census field staff, and data supervisors who collect and process this information, and the Community Liaison Group who run the community engagement programmes. This article is published with the permission of the Director of the Kenya Medical Research Institute.

