## supplementary file for "Impact of the COVID-19 epidemic on mortality in rural coastal Kenya"

**Authors and affiliations**

Otiende M^1*^, Nyaguara A^1^, Bottomley C^2^, Walumbe D^1^, Mochamah G^1^, Amadi D^1^, Nyundo C^1^, Kagucia EW^1^, PhD; Etyang AO^1^, Adetifa IMO^1,2^, Maitha E^3^, Chondo E^3^, Nzomo E^4^, Aman R^5^, Mwangangi M^5^, Amoth P^5^, Kasera K^5^, Ng’ang’a W^6^, Barasa E^1^, Tsofa B^1^, Mwangangi J^1^, Bejon P^1,7^, Agweyu A^1^, Williams TN^1,8^, Scott JAG^1,2^

^1^ KEMRI-Wellcome Research Trust Programme, Kilifi, Kenya

^2^ Department of Infectious Disease Epidemiology, London School of Hygiene & Tropical Medicine, London, UK

^3^ Department of Health, Kilifi County, Kenya

^4^ Kilifi County Hospital, Kilifi, Kenya

^5^ Ministry of Health, Government of Kenya, Nairobi, Kenya

^6^ Presidential Policy and Strategy Unit, The Presidency, Government of Kenya, Nairobi, Kenya

^7^ Nuffield Department of Clinical Medicine, University of Oxford, Oxford, UK

^8^ Imperial College, London, UK

Appendix 1

Statistical models

**Equation S1: Negative binomial model for mortality**

$$y_{t}∽Negative Binomial \left( \mu_{t},\phi\right),$$

$$\log\mu_{t}=\beta_{0}+\beta_{1}t+\beta_{2}\cos\left( \frac{2\pi t}{12} \right)+\beta_{3}\sin\left( \frac{2\pi t}{12} \right)+\log\gamma_{t}.$$

$$y_{t}=no. of deaths per month$$

The model includes terms to account for log-linear trend and seasonality (sine and cosine terms) and an offset ($\log\gamma_{t})$to account for changes in person years of observation.

**Equation S2: Local level model for all admissions**

$y_{t}=\mu_{t}+\varepsilon_{t}$, $\varepsilon_{t}∽Normal \left( 0,\sigma_{\mathcal{E}}^{2} \right),$

$\mu_{t+1}=\mu_{t}+\eta_{t}, \eta_{t}∽Normal (0,\sigma_{\eta}^{2})$.

$$y_{t}=per capita monthly \mathrm{admissions}$$

The model does not include any covariates. Instead it is assumed that the mean ($\mu_{t})$follows a random walk.

**Equation S3: Linear regression model with ARMA errors for pneumonia admissions**

$\log y_{t}=\beta_{0}+\beta_{1}t+\epsilon_{t}$.

$$y_{t}=per capita quarterly pneumonia admissions$$

The model includes a term for log-linear trend and the error ($\epsilon_{t})$follows an ARMA process. Akaike’s Information Criterion (AIC) is used to choose the order of the ARMA process.

### **Table S1**. Additional interim VA questions for identification of possible COVID-19 deaths

| **ID** | **Question** | **Responses** |
| --- | --- | --- |
| id10482 | Was there any diagnosis by a health professional of COVID-19? | yes; no; don’t know; refused to answer |
| id10483 | Did he/she have a recent test by a health professional for COVID-19? | yes; no; don’t know; refused to answer |
| id10484 | What was the result? | positive; negative; unclear; don’t know; refused to answer |
| id10485 | Did he/she suffer from extreme fatigue? | yes; no; don’t know; refused to answer |
| id10486 | Did he/she experience a new loss, change or decreased sense of smell or taste? | yes; no; don’t know; refused to answer |
| id10487 | In the two weeks before death, did he/she live with, visit, or care for someone who had any COVID-19 symptoms, or a positive COVID-19 test? | yes; no; don’t know; refused to answer |

### **Table S2.** Timeliness of ascertainment of deaths by re-enumeration round

| **Re-enumeration round (n)** | **start date** | **finish date** | **all deaths detected by date of occurrence** | **deaths detected in round**  **n** | **% detected in round**  **n** | **deaths detected in round n+1** | **% detected in round n+1** | **deaths detected after round n+1** | **%. detected after round n+1** |
| --- | --- | --- | --- | --- | --- | --- | --- | --- | --- |
| 37 | 16/11/2015 | 02/05/2016 | 611 | 400 | 65.5 | 197 | 32.2 | 14 | 2.3 |
| 38 | 03/05/2016 | 23/08/2016 | 428 | 225 | 52.6 | 198 | 46.3 | 5 | 1.2 |
| 39 | 24/08/2016 | 09/01/2017 | 440 | 207 | 47.0 | 233 | 53.0 | 0 | 0.0 |
| 40 | 10/01/2017 | 13/07/2017 | 698 | 432 | 61.9 | 265 | 38.0 | 1 | 0.1 |
| 41 | 14/07/2017 | 09/01/2018 | 661 | 327 | 49.5 | 332 | 50.2 | 2 | 0.3 |
| 42 | 10/01/2018 | 30/04/2018 | 396 | 208 | 52.5 | 184 | 46.5 | 4 | 1.0 |
| 43 | 02/05/2018 | 03/09/2018 | 471 | 238 | 50.5 | 228 | 48.4 | 5 | 1.1 |
| 44 | 04/09/2018 | 03/01/2019 | 410 | 213 | 52.0 | 196 | 47.8 | 1 | 0.2 |
| 45 | 04/01/2019 | 23/04/2019 | 356 | 174 | 48.9 | 178 | 50.0 | 4 | 1.1 |
| 46 | 24/04/2019 | 18/08/2019 | 435 | 243 | 55.9 | 181 | 41.6 | 11 | 2.5 |
| 47 | 19/08/2019 | 02/01/2020 | 442 | 223 | 50.5 | 214 | 48.4 | 5 | 1.1 |
| 48 | 03/01/2020 | 11/01/2021 | 1305 | 556 | 42.6 | 734 | 56.2 | 15 | 1.1 |
| 49 | 12/01/2021 | 03/05/2021 | 375 | 199 | 53.1 | 172 | 45.9 | 4 | 1.1 |
| 50 | 04/05/2021 | 23/09/2021 | 601 | 313 | 52.1 | 288 | 47.9 |  |  |
| 51 | 24/09/2021 | 14/01/2022 | 247 | 247 |  |  |  |  |  |
| 37-47 | 16/11/2015 | 02/01/2020 | 5348 | 2890 | 54.0 | 2406 | 45.0 | 52 | 1.0 |

Baseline ascertainment was assessed for rounds 37-47 because, in subsequent rounds, the two-round lag period required to ascertain all deaths was interrupted by the pandemic (R48-49) or has not yet been completed (R50-51). This summary is based on events captured on or before 14^th^ January 2022.

### **Table S3.** Summary of excess deaths in infants

| **Age** | **Deaths** | | **Excess mortality**^*^ | | |
| --- | --- | --- | --- | --- | --- |
|  | **observed** | **expected** | **deaths** | **%** | **95% PI** |
| *1^st^ April 2020-17^th^ February 2021* | | | | | |
| 0-28d | 66 | 89 | -23 | -25.8 | -37.5, -1.5 |
| 29-365d | 31 | 50 | -19 | -38.0 | -48.8, -16.5 |
| <1 | 97 | 139 | -42 | -30.2 | -45.8, -11.3 |
| *18^th^ February 2021-23^rd^ September 2021* | | | | | |
| 0-28d | 49 | 62 | -13 | -21.0 | -37.3, 8.6 |
| 29-365d | 34 | 34 | 0 | 0 | -25.5, 37.2 |
| <1 | 83 | 96 | -13 | -13.5 | -34.5, 7.1 |
| *1^st^ April 2020-23^rd^ September 2021* | | | | | |
| 0-28d | 115 | 150 | -35 | -23.3 | -34.8, -3.5 |
| 29-365d | 65 | 85 | -20 | -23.5 | -36.4, -3.8 |
| <1 | 180 | 235 | -55 | -23.4 | -36.5, -9.0 |

*Percentages are the proportion of excess deaths out of all expected deaths.

### **Table S4.** Summary of excess deaths between 18^th^ February 2021-23rd September 2021

| **Age** | **Deaths** | | **Excess mortality**^*^ | | |
| --- | --- | --- | --- | --- | --- |
|  | **observed** | **expected** | **deaths** | **%** | **95% PI** |
| 1-4y | 32 | 39 | -7 | -17.9 | -40.8, 13.1 |
| 5-14y | 31 | 43 | -12 | -27.9 | -44.5, -8.5 |
| 15-44y | 118 | 138 | -20 | -14.5 | -25.8, 0.1 |
| 45-64y | 162 | 147 | 15 | 10.2 | -1.2, 30.0 |
| >64y | 425 | 340 | 85 | 25.0 | 11.8, 38.6 |
| All ages | 768 | 707 | 61 | 8.6 | 1.8, 14.6 |

*Percentages are the proportion of excess deaths out of all expected deaths.

### **Table S5.** Excess deaths 1^st^ April 2020-23rd September 2021 in KHDSS residents aged ≥1y adjusted for under ascertainment after 3^rd^ May 2021

| **Age** | **Deaths** | | **Excess mortality** | | | |
| --- | --- | --- | --- | --- | --- | --- |
| **group** | **observed** | **expected** | **N** | **%** | **95% PI** | **Risk /100,000** |
| *1^st^ April 2020 - 23^rd^ September 2021 adjusted for under-ascertainment* | | | | | |  |
| 1-4y | 80.15 | 96 | -15.85 | -16.5 | -30.2, 1.1 | -43.9 |
| 5-14y | 99.20 | 111 | -11.8 | -10.6 | -25.9, 12.5 | -12.8 |
| 15-44y | 322.74 | 343 | -20.26 | -5.9 | -13.3, 5.6 | -16.3 |
| 45-64y | 372.23 | 358 | 14.23 | 4.0 | -3.5, 16.8 | 44.1 |
| ≥65y | 899.21 | 810 | 89.21 | 11.0 | 5.0, 19.8 | 699.8 |
| All ages^*^ | 1773.53 | 1718 | 55.53 | 3.2 | -0.6, 8.1 | 18.7 |

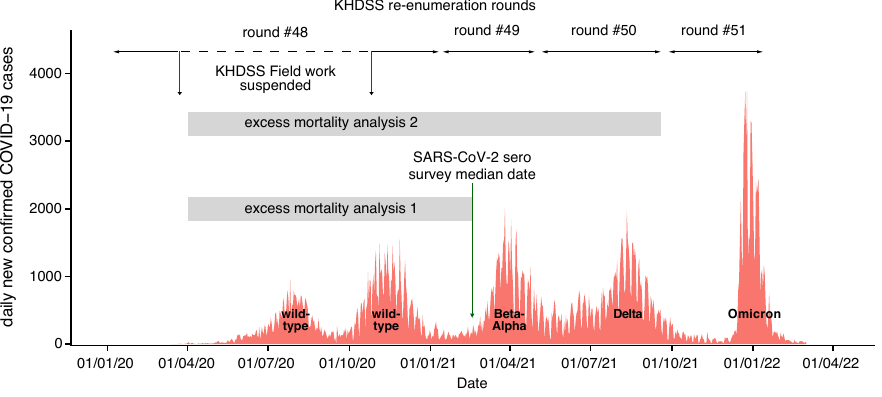

### **Figure S1.** Timeline of the 5 COVID-19 waves, KHDSS re-enumeration rounds and excess mortality analysis windows.

### Re-enumeration round 48 started on 3^rd^ January 2020 and was interrupted by the pandemic on 23^rd^ March for 7 months, resuming on 26^th^ October 2020; it was completed on 11^th^ January 2021. Round 49 spanned 12^th^ January-3rd May 2021, round 50 4^th^ May-23^rd^ September and round 51 24^th^ September 2021-14^th^ January 2022. The dotted horizontal line shows the period when there was no KHDSS fieldwork during round 48. The orange data series are the daily number of new cases of COVID-19 in Kenya. The predominant variant behind each wave is denoted at the base of each wave. Data source: COVID-19 cases were obtained from Our World in Data (https://ourworldindata.org)

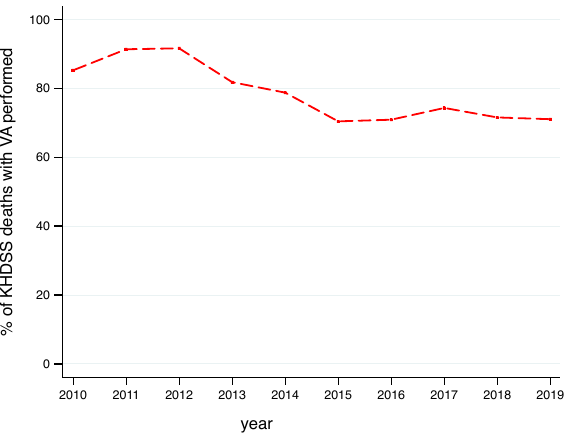

### **Figure S2.** The proportion of all deaths recorded in Kilifi Health and Demographic Surveillance System that are investigated by verbal autopsy during the baseline period 2010-2019.

### Between 1^st^ January 2020 and 3^rd^ May 2021, 1,183 (66%) of 1,796 deaths were investigated by verbal autopsy. Reasons for incomplete investigation include; inappropriate respondent, respondent not at home, postponed interview or refusal by the respondent. Given longer intervals between death and ascertainment of death in 2020/21 finding an appropriate respondent was marginally more difficult

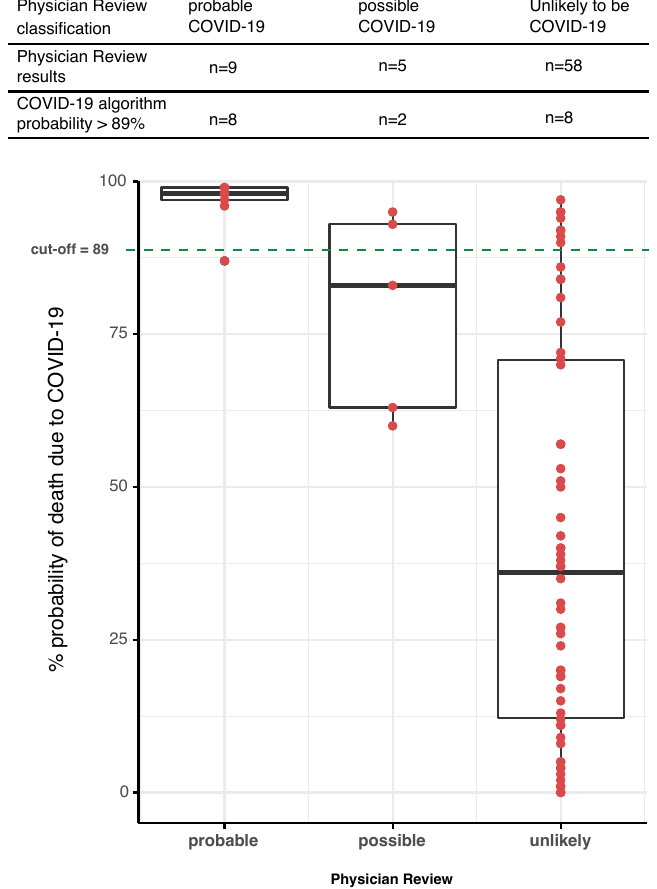

### **Figure S3.** Verbal Autopsy Assignment of COVID-19 as cause of death by CRMS algorithm and by physician review.

### Red dots are the probability values from the CRMS algorithm. They have been overlaid on their respective box plots which shows their distribution. The plots have been categorized according to the physician review categories. The green horizontal dotted line shows the probability cut-off value (89%) beyond which a death is classified as COVID-19 related by the CRMS algorithm.

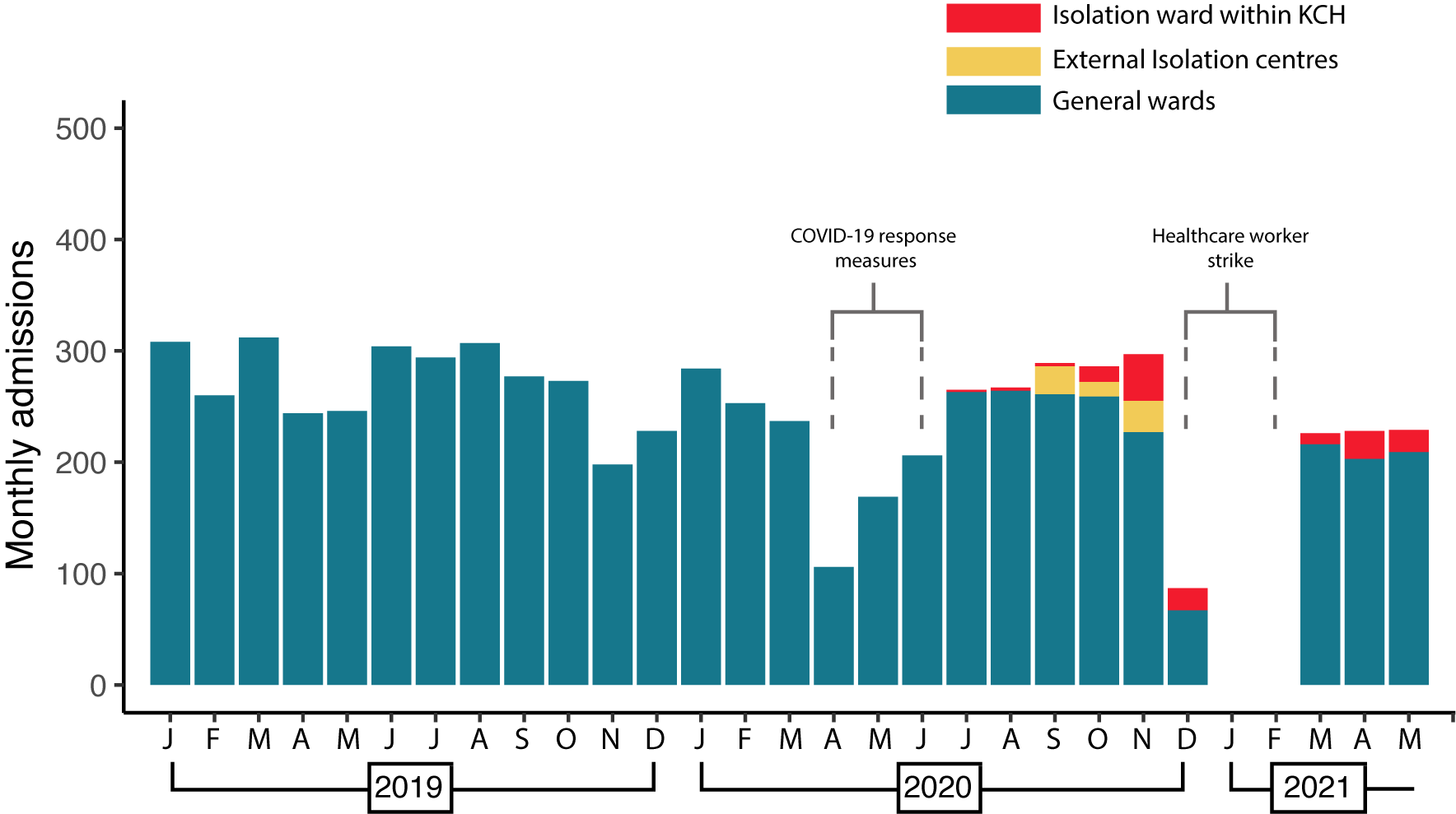

### **Figure S4.** Monthly admissions to the general medical wards in KCH, to the isolation ward within KCH and to the Kilifi County COVID-19 isolation centres among adults, aged ≥15 years.

### We were unable to incorporate in our analyses potential COVID-19 cases who presented to the other emergency response services in Kilifi County because they were not linked to KHDSS residence status. These include a 14-bed amenity ward attached to Kilifi County Hospital and two isolation centres external to the hospital. The numbers admitted in these centres were small. The admissions recorded in this figure represent all patients at all three facility types, whether or not they were residents of KHDSS. Of the total clinical burden that could be attributed to COVID-19 there is no obvious excess during the period of the pandemic.

### The decline in admissions visible between April – June 2020 was attributable to the nationwide lockdown in the first few days after the index case in Kenya. The decline and missing data in December 2020 and January-February 2021 is attributable to a healthcare workers’ strike during which none of the clinical services were in operation.

Appendix 2 Data Quality Checks

Because there is a theoretical possibility of under-ascertainment of deaths during the period 23 March 2020 to 26 October 2020, when KHDSS field operations were suspended, we performed two data quality checks to investigate this possibility.

Firstly, we examined the source of information on deaths, current residents and out-migrants. If a fieldworker cannot find a household member during a re-enumeration round, the interview may be conducted with a respondent from a neighbouring household. One possibility to explain under-ascertainment of deaths during the pandemic period is that the fieldwork during this period relied more heavily on external respondents. We looked at the proportion of reports given by a respondent who was a resident of the same household.

Table S4 illustrates the variation in this proportion over the 11-year period of the mortality analysis. Deaths were reported by a member of the same household within a range of 71-82% in the period before fieldwork was suspended by Government restrictions on 23^rd^ March 2020. After fieldwork resumed on October 26^th^ there was no evidence of an increase in unreliable, external witnesses; in fact, the reporting of deaths was more likely to originate from household members than during the baseline period, probably because of reduced mobility and a greater likelihood of finding a respondent at home

### **Table S6.** Distribution of KHDSS respondents by time period of fieldwork.

|  | % of events reported by  members within the household | | |
| --- | --- | --- | --- |
| Year | deaths | outmigrations | enumerations |
| 2010 | 82 | 87 | 85 |
| 2011 | 80 | 85 | 85 |
| 2012 | 78 | 85 | 85 |
| 2013 | 81 | 84 | 87 |
| 2014 | 72 | 84 | 86 |
| 2015 | 72 | 84 | 86 |
| 2016 | 73 | 90 | 86 |
| 2017 | 80 | 87 | 87 |
| 2018 | 75 | 85 | 86 |
| 2019 | 71 | 86 | 86 |
| 2020 Jan-Mar | 73 | 87 | 87 |
| 2020 Oct-2021 May | 85 | 81 | 85 |

Secondly, we explored the possibility that travel restrictions may have reduced a phenomenon known as the unhealthy in-migrant; this is a scenario where a sick person returns to his or her rural home to die. This contributes to all-cause mortality in KHDSS but such sick individuals, about to die, may not have been able to travel to Kilifi during the pandemic period. If this effect was substantial, the unhealthy in-migrant deaths unobserved during the pandemic period may have masked an excess of COVID-19 related deaths among stable residents of KHDSS. To explore this potential bias we calculated the 12-month risk of death in a cohort of residents selected on 23^rd^ March 2020, the day after the first case of COVID-19 was detected in Kilifi, and compared this cohort to similar cohorts selected on the same date in previous years. Table S5 shows the annual mortality risks. Figure S6 shows the survival of each cohort from 2010-2020. These analyses illustrate the mortality risk and survival function after excluding any in-migrants during the risk period. The results do not suggest an excess of mortality risk or reduced survival in 2020/21 among stable residents of the KHDSS.

### **Table S7.** Age-adjusted annual risk of death in cohorts of residents selected on 23 March of each year from 2010-2020

| **Cohort year** | **All ages** | | **≥15 years** | |
| --- | --- | --- | --- | --- |
|  | **Population** | **% mortality within 1 year** | **Population** | **% mortality within 1 year** |
| 2010 | 248,184 | 0.60 | 127,277 | 0.95 |
| 2011 | 253,947 | 0.53 | 130,163 | 0.83 |
| 2012 | 258,800 | 0.52 | 133,478 | 0.85 |
| 2013 | 266,915 | 0.51 | 139,089 | 0.82 |
| 2014 | 271,337 | 0.50 | 142,547 | 0.76 |
| 2015 | 275,636 | 0.52 | 146,183 | 0.82 |
| 2016 | 278,677 | 0.46 | 149,626 | 0.74 |
| 2017 | 285,120 | 0.52 | 154,094 | 0.83 |
| 2018 | 289,940 | 0.44 | 157,796 | 0.68 |
| 2019 | 296,084 | 0.42 | 162,591 | 0.64 |
| 2020 | 300,742 | 0.44 | 166,817 | 0.68 |

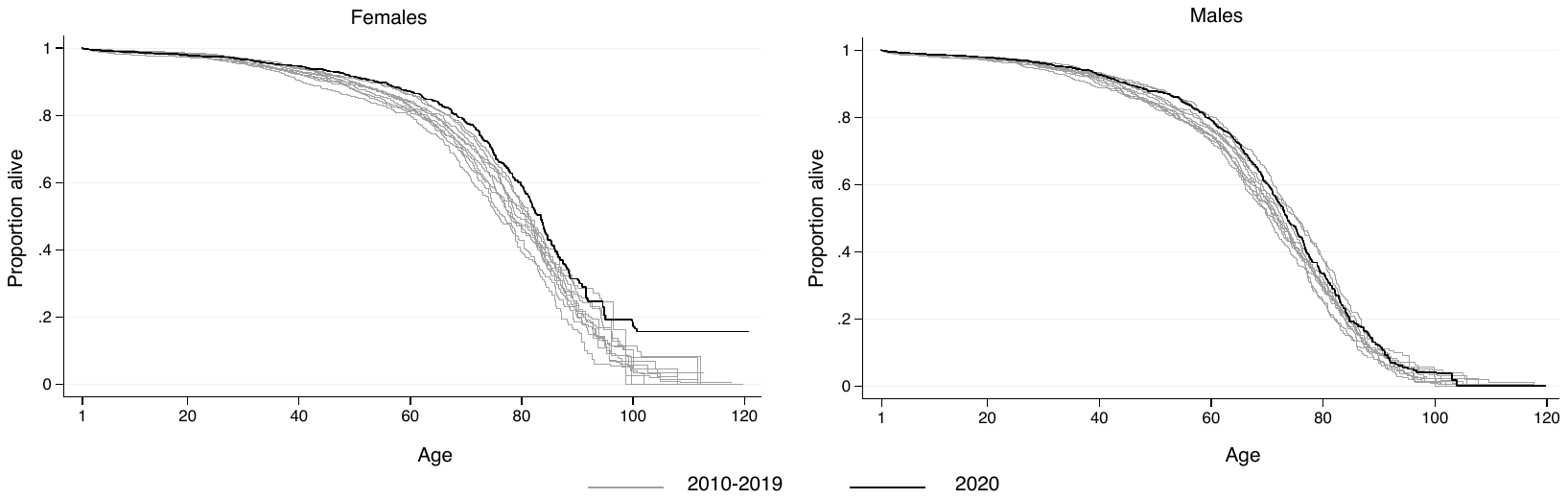

### **Figure S5**. Annual period survival curves for cohorts resident on the 23^rd^ March each year from 2010-2020 for males and females.

### Survival time is age, and the survival function starts at age 1 year, for consistency with other analyses. The light grey lines are individual survival curves of cohorts for the years 2010-2019 and the black solid lines are the survival curves for the 2020 cohort.
